# Temporal dynamics of skin microbiota and immune correlates in psoriasis patients receiving systemic treatment

**DOI:** 10.1101/2023.11.27.23298999

**Authors:** Su-Hsun Liu, Yu-Huei Huang, Hao-Jui Weng, Tsen-Fang Tsai, Huang-Yu Yang, Leslie Y Chen, Yen-Ling Chiu, Hsiao-Yun Yu, Yi-Chieh Chiu, Chao-Yu Ng, Ya-Ching Chang, Chung-Yee R Hui, Yhu-Chering Huang

**Affiliations:** International Health Program, National Yang Ming University, Taipei, Taiwan; Health Management Center, Far Eastern Memorial Hospital, New Taipei City, Taiwan; College of Medicine, Chang Gung University, Taoyuan City, Taiwan; Department of Dermatology, Chang Gung Memorial Hospital, Taoyuan, Taiwan; Department of Dermatology, National Taiwan University Hospital, Taipei, Taiwan; Department of Dermatology, School of Medicine, College of Medicine, Taipei Medical University, Taipei, Taiwan; Department of Dermatology, Taipei Medical University-Shuang Ho Hospital, New Taipei city, Taiwan; Department of Dermatology, School of Medicine, College of Medicine, National Taiwan University, Taipei, Taiwan; Department of Nephrology, Chang Gung Memorial Hospital, Taoyuan, Taiwan; Department of Medical Research, Chang Gung Memorial Hospital, Taoyuan, Taiwan; Department of Nephrology, Far Eastern Memorial Hospital, New Taipei City, Taiwan; Department of Pediatrics, Chang Gung Memorial Hospital, Taoyuan, Taiwan

**Keywords:** Skin microbiota (Microbiology), psoriasis, clinical outcomes, epidemiology (Cohort study), biomarkers, systemic treatment

## Abstract

**Background:** How skin microbiota in psoriasis patients responded to systematic therapeutics remained unknown.

**Objectives:** To profile temporal shifts in transcriptionally active skin microbiota in psoriasis patients receiving systemic therapies.

**Methods:** We prospectively enrolled 61 psoriasis patients and 29 skin-healthy controls in 2015-2019. Using RNA-based 16S rRNA gene sequencing, we analyzed 969 samples from skin lesions and compared microbial abundance and diversity by therapeutic classes and disease severity.

**Results:** Lesional microbiota in patients on conventional systemics and TNF-*α* inhibitor was different in relative abundances in Firmicutes (7.83% higher, adjusted P < 0.001) and Proteobacteria (6.98% lower, adjusted P < 0.01) from that in patients on anti-interleukin monoclonal antibodies (anti-ILAb) at baseline. The only difference during treatment was a 1.47% lower abundance in Bacteroides associated with nonbiologics use (adjusted P < 0.01). We identified no indicator taxa by disease severity at baseline yet noticed that a minor relative reduction in *Corynebacterium* sp. was associated with clinical responses to treatment.

Compared to anti-ILAb, TNF-*α* inhibitor and nonbiologics were associated with -0.21 lower Shannon Diversity (adjusted P < 0.01) and 0.03 higher Shannon Evenness (adjusted P < 0.01). Results of ordinated principal coordinates analysis revealed that, lesional microbiota from patients of these 3 therapeutic groups was compositionally distinct. Our work also demonstrated concurrent changes in clonal shifts in systemic T cell receptor clonotypes that were associated with systemic use of biologics.

**Conclusions:** Community abundances and diversities of skin microbiota may be useful in distinguishing skin microbiota from patients receiving different systemic therapeutics. Specifically, use of anti-ILAb and TNF-*α* inhibitor was associated with sample-wise microbial abundances and diversities, but not richness, over time. These findings highlighted the potential utility of skin microbiota as biomarkers for personalized treatment plans in patients with moderate-to-severe psoriasis.

## INTRODUCTION

Psoriasis vulgaris (psoriasis) is a chronic immune disease that manifests as well-defined scaly skin plaques, which may be associated with arthritis and systemic comorbidities [1]. Both innate immunity and T-cell response contribute to the disease pathogenesis, constituting major therapeutic targets for effective systemic therapies [2]. Recently, skin microbiota has emerged as novel candidate biomarkers for disease management [3, 4].

Despite the proven clinical effectiveness of anti-psoriatic therapies, few studies have examined therapy-specific influences on lesional microbiota longitudinally [5]. A recently-updated Cochrane review showed that biologics, including TNF-*α* inhibitor and anti-ILAb, were more effective in achieving clinical remission as measured by PASI 90 (90% reduction in Psoriasis Area and Severity Index relative to the baseline) than the small molecules and conventional systemics in the trial setting [2]. However, whether and how conventional therapies, TNF-*α* inhibitor, and anti-ILAb differentially alter the host microbiota in a non-trial setting remains unstudied. Besides, whether clinical responses were associated with any shifts in lesional microbiota has not been well-characterized [3]. As such, we sought to delineate relationships among systemic therapies, treatment responses, and characteristics of lesional microbiota in psoriasis patients via serial sampling. Particularly, to reduce environmental influences on the low biomass of skin samples [6] and to provide an in-depth insight into the relevant responses to a newly-prescribed regimens [7, 8], we sought to profile transcriptionally-active skin microbiota using bacterial 16S rRNA transcript sequencing. We also explored temporal changes in T cell receptor (TCR) repertoire diversity during treatment for a better understanding of treatment effects on the host immunity.

## MATERIALS AND METHODS

### Subject enrollment

Between May 2015 and November 2019, we enrolled and followed adults (aged 20 years or older) with psoriasis at Dermatology Outpatient Clinics of two tertiary hospitals (Chang Gung Memorial Hospital and National Taiwan University Hospital) in Taiwan. Patients were eligible for study entry when initiating new systemic therapy; majority of the washout period was > 4 weeks. We excluded those with a recent exposure to topical or systemic antibiotics, immunosuppressants, or chemotherapies within 12 weeks prior to study enrollment. In 2016- 2019, we also recruited skin-healthy adults who did not have diagnoses of chronic inflammatory skin disorders but either visited the same clinics for mole excision or alopecia treatment, or were care givers of the index patients. Center-specific Institution Review Boards reviewed and approved the study protocols, separately (CGMH 103-6837B, 104-9746B, 201702308B0C501; NTUH 201809061RINB).

### Data and sample collection

After obtaining written informed consent, well-trained research assistants conducted in-person interviews to collect sociodemographic information, medical history, and behavioral habits.

Patients also reported quality of life and anxiety/depressive symptoms using Dermatology Life Quality Index (DLQI)[9] questionnaire and Center for Epidemiologic Studies Depression Scale (CESD-S)[10] at baseline and follow-up visits. According to European S3-Guidelines [11], we grouped patients by non-biologics (acitretin, cyclosporin, methotrexate); anti-interleukin monoclonal antibodies (anti-ILAb: ustekinumab, secukinumab, ixekizumab, and guselkumab); tumor necrosis factor-*α* antagonists (TNF-*α* inhibitor: adalimumab, golimumab, and etanercept). Treatment responses were absolute PASI scores and PASI 75 responses.

To profile skin microbiota from sequencing 16S rRNA gene transcripts, we used Tween20- moistened FLOQSwabs (Copan, Murrieta, CA, USA) to collect scrapes over an area of 2 x 2 cm^2^ on pre-specified surfaces with active lesions [12, 13], including bilateral scalp, post-auricular hairlines, extensor forearms, pretibial shafts, and palms; these sampling sites also applied to skin-healthy controls. To facilitate swab collection, we requested the study participants to refrain from bathing or body scrubbing at least 24 hours prior to a study visit. We also encouraged each participant to cover up sampling sites with cotton-made clothing to reduce environmental influences while in the study.

### 16S rRNA gene transcript isolation, amplification, sequencing, and processing

We performed RNA isolation by firstly disrupting bacterial cell walls in a chemical lysis buffer [7]. Reverse transcription of total RNA immediately followed the recovery of RNA samples using the SuperScript RT-PCR kit (Invitrogen, Carlsbad, CA, USA) according to the manufacturer’s manual. For each PCR sample, we confirmed the presence of amplicons by gel electrophoresis. We included 2-3 negative controls for each library preparation as suggested in the literature [14, 15], and mock community DNA (Zymo Research, Irvine, CA, USA) as positive controls [16].

We applied ‘mothur’ (version 1.43.0) to the raw sequence reads using a modified reference region for sequence alignment [17, 18] and clustered the filtered reads into operational taxonomy units (OTU) at 97% identify before subsequent statistical analysis and graphing in R (v 4.0.2) [19]. Using R packages ‘decontam’ and ‘phyloseq’[15, 20], we identified and removed potential contaminant taxa inferred from 63 negative control samples [21], singletons, and samples with low coverage (< 400 reads/ sample). In total, we included 969 samples (82.0% of sequenced samples) with a median of 43 577 reads per sample (interquartile range [IQR]: 33 987-55 972) in the following analysis.

### TCR repertoire profiling

We collected peripheral blood samples in EDTA tubes from 9 anti-ILAb users before and 12 weeks after treatment began. We also collected 1 blood sample from 7 skin-healthy participant at baseline. Using flow cytometry, we sorted CD3+ T cells, from which RNA isolation was performed by employing RNeasy Mini Kit (Qiagen). Procedures of the library preparation included nested amplifications targeting the V-C regions of the complementarity-determining region (CDR3) of TCR using TCR*αβ* kits (iRepertoire, Huntsville, AL, USA).

### Statistical Analysis

#### Patient data

We described and compared baseline participants’ characteristics by disease status and therapy class (patients only) using median tests for independent continuous variables and Chi-squared or Fisher’s exact test for categorical ones. We tested our hypothesis using multivariable quantile regression analysis with robust or bootstrapped (3 000 replicates) variance estimation to account for repeated measures. All longitudinal data analysis considered observations over the 12-15 weeks of follow-up period to allow for sufficient statistical power.

We used Stata-MP (Version 13.1) with a 2-tailed significance of 0.01 for comparative analysis [22], and employed R packages ‘ggplot2’ and ‘vegan’ for graphics production [23, 24].

#### Microbiota data

We calculated *α*-diversity measures of richness (observed number of unique OTUs, Chao1 index), Shannon diversity (ShannonD) and Shannon evenness (ShannonE) indices for quantifying within-sample variation. Based on non-metric and linear R^2^ values, we chose Bray-Curtis dissimilarity index to compare between-sample distance matrices [25]. We performed hypothesis testing via permutational multivariate analysis of variance (PERMANOVA, 10 000 permutations) [26]. We also repeated relevant analysis using read-based rarefied datasets; the results did not alter our conclusions.

#### TCR profiling

We performed descriptive analysis by sample and patient characteristics; compared differences in the normalized usage of V-and J-segment of the CDR3 gene, and in diversity metrics of CDR3 clonotypes using bootstrapped quantile regression models to estimate the empirical variance (3 000 replicates) in Stata-MP (Version 13.1) [22]. We applied a 2-tailed significance level of 0.05 for exploratory analysis on gene expression and TCR sequence data.

### Data Availability

We have deposited raw sequences of 16S rRNA transcripts and TCR transcripts to the Short Read Archive (SRA) database under the BioProject accession number PRJNA662768.

## RESULTS

### Study population

We prospectively enrolled and followed 119 participants for 12 weeks or longer between 2015 and 2019. Ten patients received an extended observation for additional 6 months while receiving maintenance biologics therapy (Fig. S1). We included 743 and 226 quality swabs from 61 psoriasis patients and 29 controls who returned at least once within 16 weeks in the current analysis (follow-up rate: 87% and 83%). The patient population comprised mainly of men (87%) (median age: 43 years) with an average disease duration of 23 years (IQR: 19-33). There were no statistical differences in patient characteristics associated with therapeutic classes at study entry. In total, 79% of patients had moderate-to-severe psoriasis; all patients of the 3 therapeutic groups began with comparable disease severity at study entry (Table 1). Patients excluded based on low-quality sequencing samples (N=5) were comparable to those included in the current analysis (Table S1).

**Table 1.** Characteristics of psoriasis patients and skin-healthy control participants at baseline, 2015-2019.

| Characteristics, % | Psoriasis Patients |  |  | P-value<sup>a</sup> | Skin-healthy Controls | P-value<sup>b</sup> |
| --- | --- | --- | --- | --- | --- | --- |
| | Non-biologics (N=6) | TNF-α Inhibitors (N=13) | Anti-ILAbs (N=42) | | (N=29) | |
| Follow-up time (days), Median (IQR) | 98 (77-112) | 103 (94-119) | 108 (84-112) | 0.318 | 84 (84-91) | <0.001 |
| Age (years), Median (IQR) | 41 (37-44) | 40 (32-58) | 44 (35-52) | 0.318 | 51 (45-58) | 0.006 |
| 20-36 | 17% | 46% | 29% | 0.189 | 3% | 0.003 |
| 36-50 | 83% | 23% | 43% |  | 45% |  |
| $\geq 50$ | 0 | 31% | 29% | | 52% | |
| Male | 100% | 92% | 83% | 0.587 | 62% | 0.007 |
| BMI (Kg/m<sup>2</sup>), Median (IQR) | 27.3 (26.8-33.0) | 28.7 (26.3-30.7) | 27.7 (25.9-29.7) | 0.450 | 24.7 (23.1-26.2) | <0.001 |
| WC (cm)<sup>c</sup>, Median (IQR) | 96 (91-102) | 97 (93-108) | 92 (82-100) | 0.240 | 79 (76-85) | <0.001 |
| Education<sup>c</sup> |  |  |  |  |  |  |
| Less than high school | 0 | 23% | 36% | 0.183 | 17% | 0.421 |
| High or vocational school | 43% | 23% | 38% |  | 34% |  |
| College or higher | 57% | 54% | 26% |  | 45% |  |
| Disease duration (years), Median (IQR) | 22 (20-36) | 21 (20-33) | 26 (19-33) | 0.422 | - | - |
| Treatment-naïve | 57% | 23% | 29% | 0.546 | - | - |
| Regular alcohol use | 14% | 23% | 24% | 0.545 | 21% | 0.946 |
| Regular smoking |  |  |  |  | N=28 |  |
| Never | 29% | 77% | 55% | 0.284 | 79% | 0.040 |
| Quitted | 0 | 0 | 12% |  | 10% |  |
| Yes | 71% | 23% | 33% |  | 10% |  |
| Regular use of antiseptics for |  |  |  |  |  |  |
| Face<sup>d</sup> | 14% | 8% | 2% | 0.226 | 0 | 0.551 |
| Hand<sup>e</sup> | 0 | 15% | 7% | 0.755 | 7% | 1.000 |
| Staphylococcal colonization |  |  |  |  |  |  |
| Lesion/ skin | 57% | 23% | 31% | 0.507 | 7% | 0.015 |
| Nares | 43% | 15% | 19% | 0.211 | 7% | 0.130 |
| Clinical Outcomes |  |  |  |  |  |  |
| PASI, Median (IQR) | 15 (14-20) | 13 (11-17) | 15 (11-24) | 0.698<sup>a</sup> | - |  |
| DLQI, Median (IQR) | 12 (8-22) | 9 (4-14) | 10 (3-15) | 0.796<sup>a</sup> | - |  |
| CESD, Median (IQR) | 17 (12-24) | 13 (11-20) | 16 (13-22) | 0.431<sup>a</sup> | 13 (12-15) | 0.059 |

Abbreviations: BMI, body mass index; CESD, Center for Epidemiologic Studies Depression Scale; DLQI, Dermatology Life of Quality Index; anti-IL-Abs, anti-interleukin monoclonal antibodies; IQR, interquartile range; PASI, psoriasis area severity index; WC, waist circumference
a. P-value for median test; Chi-square test, or Fisher's exact test among psoriasis patients using different treatment regimens
b. P-value for median test; Chi-square test, or Fisher's exact test between psoriasis patients and control subjects
c. One healthy control had missing data
d. Three control subjects had missing data
e. Three control and 1 psoriasis participants had missing data

At 12-15 weeks after treatment began, disease activity in all patients declined significantly, with the median PASI score decreasing from 15 (IQR: 11-20) to 4 (IQR: 1-8) (adjusted P for trend < 0.001). Patients in each therapeutic class had comparable trends in disease activities over time; there were no significant differential group effects (all P for interaction tests > 0.01). Self-reported DLQI also decreased substantially from 9 (IQR: 4-15) at baseline to 2 (0-8, adjusted P for trend < 0.001) whereas self-reported anxiety symptoms as quantified by CESD scores did not vary over time (Table S2).

### Normal versus lesional microbiota

#### Microbial abundances

At study entry, phylum abundances in normal skin microbiota were statistically comparable to those in lesional microbiota with or without accounting for participants’ age, sex, absence of prior treatment, and year of enrollment (Table 2). A relative dominance of Proteobacteria in lesional microbiota by 3.35% (95%CI: 1.44%, 5.25%) emerged over time (adjusted P < 0.001); simultaneous comparisons across disease status and sampling sites revealed similar results (adjusted P=0.002, Fig. S2a). Likewise, we examined abundances of the most common 6 genera as suggested in the literature [27] (Fig. S2b), and found no evidence for indicator taxa between lesional and normal skin (Table S3).

**Table 2.** Differences in phylum-level relative abundance between lesional and healthy skin microbiota at baseline and during 12-15 weeks of systemic treatment in adult psoriasis patients overall, by sex and absence of prior treatment, 2016-2019^a^.

| Relative<br>Abundances (%) | Baseline (Cross-sectional model) |  | Baseline to 12-15 wk (Longitudinal model) |  |
| --- | --- | --- | --- | --- |
|  | Unadjusted | Adjusted<sup>a</sup> | Unadjusted | Adjusted<sup>a</sup> |
| **Overall** | beta (95%CI) | beta (95%CI) | beta (95%CI) | beta (95%CI) |
| Proteobacteria | 0.03 (-0.00, 0.06) | 4.54 (0.33, 8.75)b | **2.35 (0.58, 4.11)c** | **3.35 (1.44, 5.25)d** |
| Firmicutes | -0.01 (-0.03, 0.02) | -0.11 (-2.09, 1.88) | -0.78 (2.14, 0.58) | -0.24 (-1.91, 1.42) |
| Actinobacteria | -0.01 (-0.02, -0.00)b | -1.15 (-2.73, 0.44) | -1.07 (-1.97, -0.18)b | -0.94 (-2.11, 0.23) |
| Bacteroidetes | -0.002 (-0.01, 0.01) | -0.55 (-2.20, 1.10) | -0.32 (-1.06, 0.43) | -0.15 (-1.49, 0.59) |
| **Men** |  |  |  |  |
| Proteobacteria | 4.28 (0.89, 7.67)b | 5.30 (0.24, 10.4)b | **2.66 (0.64, 4.68)c** | **4.13 (1.88, 6.38)d** |
| Firmicutes | -1.12 (-3.90, 1.65) | 0.04 (-2.13, 2.21) | -0.72 (-2.45, 1.01) | -0.97 (-2.43, 0.48) |
| Actinobacteria | -1.33 (-2.70, 0.04) | -0.88 (-2.71, 0.96) | **-1.64 (-2.71, -0.57)c** | -0.87 (-2.18, 0.45) |
| Bacteroidetes | -0.25 (-1.78, 1.29) | -0.49 (-2.33, 1.36) | -0.44 (-1.43, 0.55) | -0.19 (-1.46, 1.08) |
| **Women** |  |  |  |  |
| Proteobacteria | 0.97 (-5.71, 7.65) | -4.61 (-14.4, 5.19) | 0.53 (-3.78, 4.85) | -0.64 (-6.77, 5.49) |
| Firmicutes | -0.08 (-5.81, 5.66) | 5.08 (-5.54, 15.7) | -2.15 (-4.95, 0.64) | -0.23 (-3.99, 3.53) |
| Actinobacteria | -1.00 (-3.31, 1.32) | -0.88 (-4.47, 2.70) | -0.63 (-2.20, 0.93) | -0.97 (-2.82, 0.87) |
| Bacteroidetes | 0.16 (-2.39, 2.70) | -0.10 (-4.08, 4.06) | 0.16 (-1.15, 1.47) | -0.16 (-1.84, 1.53) |
| **With prior treatment** |  |  |  |  |
| Proteobacteria | 1.55 (-2.26, 5.36) | -0.12 (-5.69, 5.45) | **3.41 (1.67, 5.15)d** | 2.37 (-0.34, 5.08) |
| Firmicutes | 2.16 (0.29, 4.03)b | 1.16 (-1.91, 4.24) | 0.57 (-0.82, 1.96) | -0.22 (-2.07, 1.64) |
| Actinobacteria | -0.89 (-2.53, 0.74) | -2.03 (-3.92, -0.13)b | -0.98 (-2.50, 0.54) | -1.41 (-2.99, 0.16) |
| Bacteroidetes | 1.32 (0.21, 2.42) | 0.47 (-1.38, 2.32) | 0.42 (-0.70, 1.54) | 0.24 (-1.07, 1.56) |
| **Without prior treatment** |  |  |  |  |
| Proteobacteria | 4.45 (-0.14, 9.04) | 4.64 (1.05, 8.22)c | 3.39 (0.76, 6.03)b | 3.45 (-0.02, 6.92) |
| Firmicutes | **-7.38 (-10.6, -4.11)d** | 0.00 (-4.96, 4.96) | **-7.30 (-9.39, -5.21)d** | -0.04 (-3.40, 3.32) |
| Actinobacteria | 0.87 (-1.05, 2.79) | -1.14 (-3.22, 0.93) | 1.03 (-0.26, 2.33) | -1.79 (-3.59, 0.01) |
| Bacteroidetes | -0.95 (-3.39, 1.49) | -1.85 (-4.63, 0.93) | -0.03 (-1.24, 1.19) | -0.57 (-2.13, 0.98) |

Abbreviations: beta, beta coefficient estimate in median regression models; CI, confidence interval for beta coefficient
a. Age-, sex-, absence of prior treatment, study year-adjusted in baseline models; follow-up duration was also accounted for in the longitudinal models; all comparisons excluded data from 2015-2016, during which no control subjects were enrolled
b. $P < 0.05$
c. $P \leq 0.01$
d. $P \leq 0.001$

Despite an over-representation of men in the patient group, results of sex-specific comparisons between lesional and normal microbiota were alike at baseline but not longitudinally, during which a relative over-abundance of Proteobacteria was significant in samples from male patients by 4.13% (95%CI: 1.88%, 6.38%) as compared to normal skin (Table 2). Formal tests for disease status-by-sex interaction were nonsignificant. Additional stratified analysis by the absence of prior treatment found no significant subgroup effects either.

#### Microbial alpha-& beta-diversity

When compared to healthy skin, psoriatic lesions had resembling microbial richness and diversity before and during treatment; only the degree of community evenness (all unique taxa showing equal abundances) was lower in lesional than normal microbiota (adjusted P=0.01; Table S4). Correspondingly, results of Bray-Curtis distance-based PCoA showed an overall community similarity between the two groups (PERMANOVA P > 0.05, Fig. S2c).

### Lesional microbiota, clinical treatment, and responses

#### Microbial abundances

Before initiating new therapies, lesional microbiota of patients using non-biologics had 7.83% more Firmicutes (95%CI: 4.26%, 10.5%) than those using anti-ILAb (adjusted P< 0.001) whereas TNF-*α* inhibitor was associated with a 6.98% lower abundance (95%CI: -12.2%, -1.73%) in Proteobacteria than anti-ILAb (Table 3). Over time, psoriatic lesions in non-biologics users showed a relative 1.47% reduction (95%CI: -2.49%, -0.45%) in Bacteroidetes as compared to anti-ILAb users while losing the dominance of Firmicutes (adjusted P> 0.01, Fig. S3). Abundances in the four major phyla did not differ by initial disease severity or patients’ clinical responses as indicated by PASI 75 at follow-up (Table 3).

**Table 3.** Adjusted associations between phylum-level relative abundances of lesional microbiota and therapeutic class or clinical outcome at baseline and over time in psoriasis patients, 2015-2019^a^.

| Relative Abundance (%) | Therapeutic Class |  | Clinical Assessment |
| --- | --- | --- | --- |
|  | TNF-a | Nonbiologics | Severity (PASI > 10) |
| **Baseline** | beta (95% CI) | beta (95% CI) | beta (95% CI) |
| Proteobacteria | **-6.98 (-12.2, -1.73)b** | 0.62 (-4.04, 5.28) | -1.48 (-4.23, 1.26) |
| Firmicutes | 1.27 (-2.57, 5.11) | **7.83 (4.26, 10.5)d** | 0.93 (-1.49, 3.35) |
| Actinobacteria | -0.63 (-1.79, 0.52) | -0.53 (-1.52, 2.59) | -0.60 (-1.93, 0.72) |
| Bacteroidetes | 0.46 (-2.45, 3.37) | -2.13 (-3.99, -0.27)c | -0.76 (-2.86, 1.34) |
| **Baseline to 12-15 wk** |  |  | Response (PASI 75) |
| Proteobacteria | -1.67 (-4.40, 1.06) | 0.84 (-1.94, 3.62) | -1.08 (-3.51, 1.35) |
| Firmicutes | 0.10 (-2.06, 2.26) | 4.15 (0.79, 7.52)c | 0.30 (-1.73, 2.32) |
| Actinobacteria | -0.73 (-1.50, 0.03) | 0.65 (-0.73, 2.03) | -0.75 (-1.68, 0.19) |
| Bacteroidetes | 0.13 (-1.45, 1.71) | **-1.47 (-2.49, -0.45)b** | -0.50 (-1.68, 0.69) |

Abbreviations: Anti-ILAbs, anti-interleukin monoclonal antibodies; beta, beta coefficient estimate in median regression models; CI, confidence interval for beta coefficient; PASI, psoriasis area severity index
a. Age-, sex-, absence of prior treatment, study year, and follow-up duration-adjusted quantile regression models; baseline PASI scores were also adjusted in evaluating associations with clinical responses; reference group was patients using anti-ILAbs in comparisons by therapeutic class
b. $P < 0.01$
c. $P < 0.05$
d. $P \leq 0.001$

Stratified analysis by prior treatment history revealed a higher abundance of Firmicutes in nonbiologics-associated samples from patients with previous treatment history (6.93%, 95%CI: 5.84%, 8.02%) but a lack thereof in those who were treatment-naïve. Similarly, subgroup analysis by sex showed a significant reduction in Proteobacteria (-17.5%, 95%CI: -29.0%, -6.1%) in women using TNF-*α* inhibitor as compared to anti-ILAb but no statistical differences by therapy group in men.

At genus levels, *Staphylococcus* sp. was more abundant in samples from the non-biologics group than the anti-ILAb group (adjusted P< 0.001) at baseline. There were no significant differential abundances by baseline disease severity (PASI > 10). Longitudinally, classes of systemic therapies were not associated with any indicator genera considered. Instead, lesions of PASI 75 responders had a 0.38% decrease in *Corynebacterium* sp. but nonsignificant differences in other genera relative to poor responders (Table S5).

#### Microbial alpha-& beta-diversity

Before treatment began, *α*-diversity measures of skin microbiota appeared comparable among samples from the three therapeutic groups, or by initial degrees of disease severity (Table 4). During the follow-up period, we noted a significant decrease (-0.21; 95%CI: -0.37, -0.06) and increase (0.03; 95%CI: 0.01, 0.06) in Shannon D and Shannon E of the TNF-*α* inhibitor group and the non-biologics group relative to the reference group, separately (Fig. 1a). Results of distance-based analysis of variance suggested significant pair-wise dissimilarities between samples from healthy skin microbiota and lesional microbiota of patients receiving anti-ILAb or TNF-*α* inhibitor (both permuted P < 0.001, Fig. 1b). On the other hand, samples from non-biologics users and healthy controls were compositionally similar as shown in unconstrained PCoA (Fig. S4a). Again, sample-wise *α*-diversity measures in PASI 75 responders and poor responders were indistinguishable during treatment (Table 4), wherefore these results were compatible to results of *β*-diversity analysis (Fig. S4b).

**Fig 1a.**
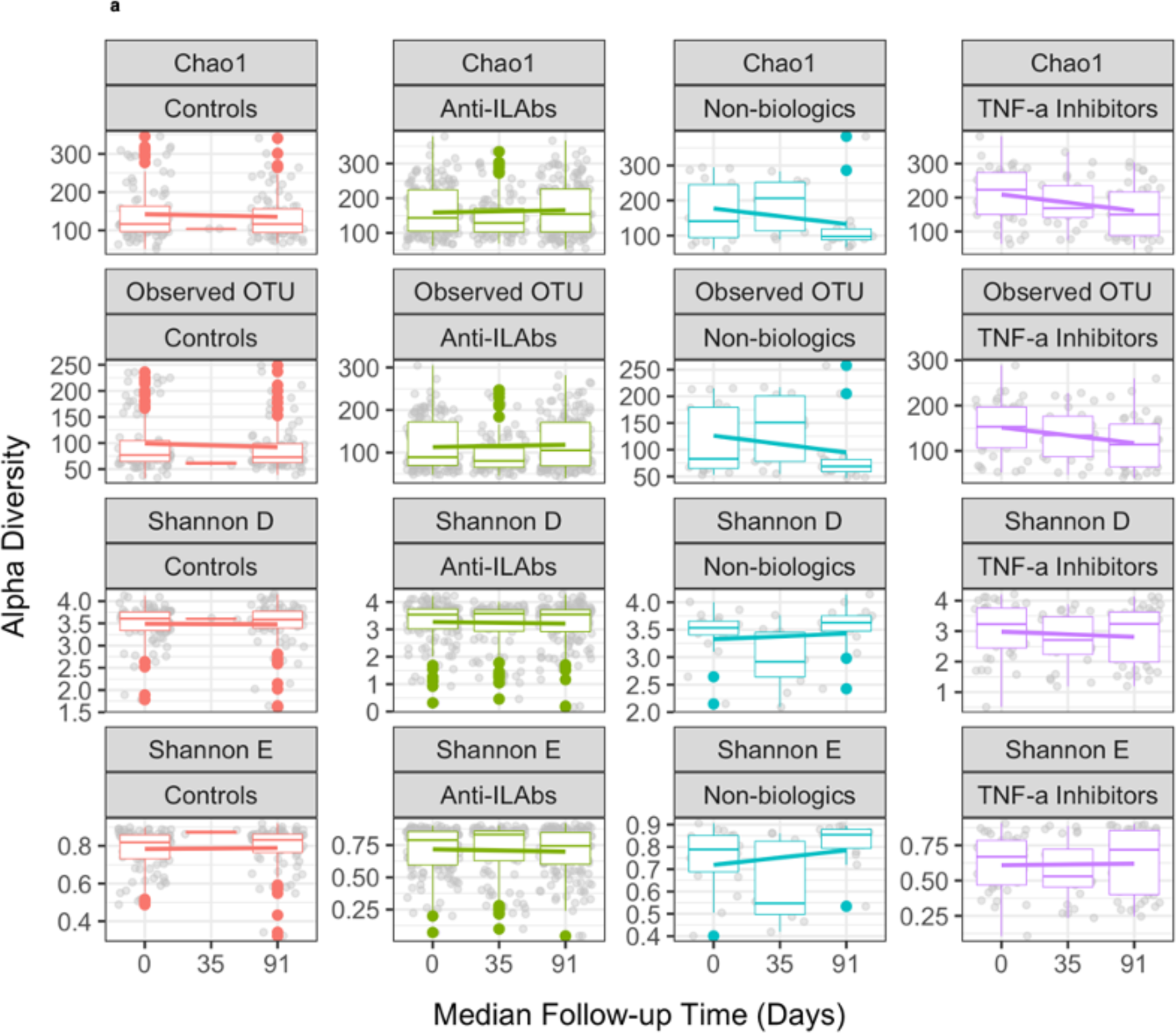
Alpha diversity measures for quantifying within-sample microbial richness (OTU, Chao1), diversity (Shannon D), and compositional evenness (Shannon E) in samples from healthy controls and psoriasis patients in the 3 therapeutic classes. Over the study period, significant differences included a relative decrease and increase in Shannon D and Shannon E in samples associated with TNF-*α* Inhibitor and nonbiologics as compared to anti-ILAb (adjusted P<0.01).

**Fig 1b.**
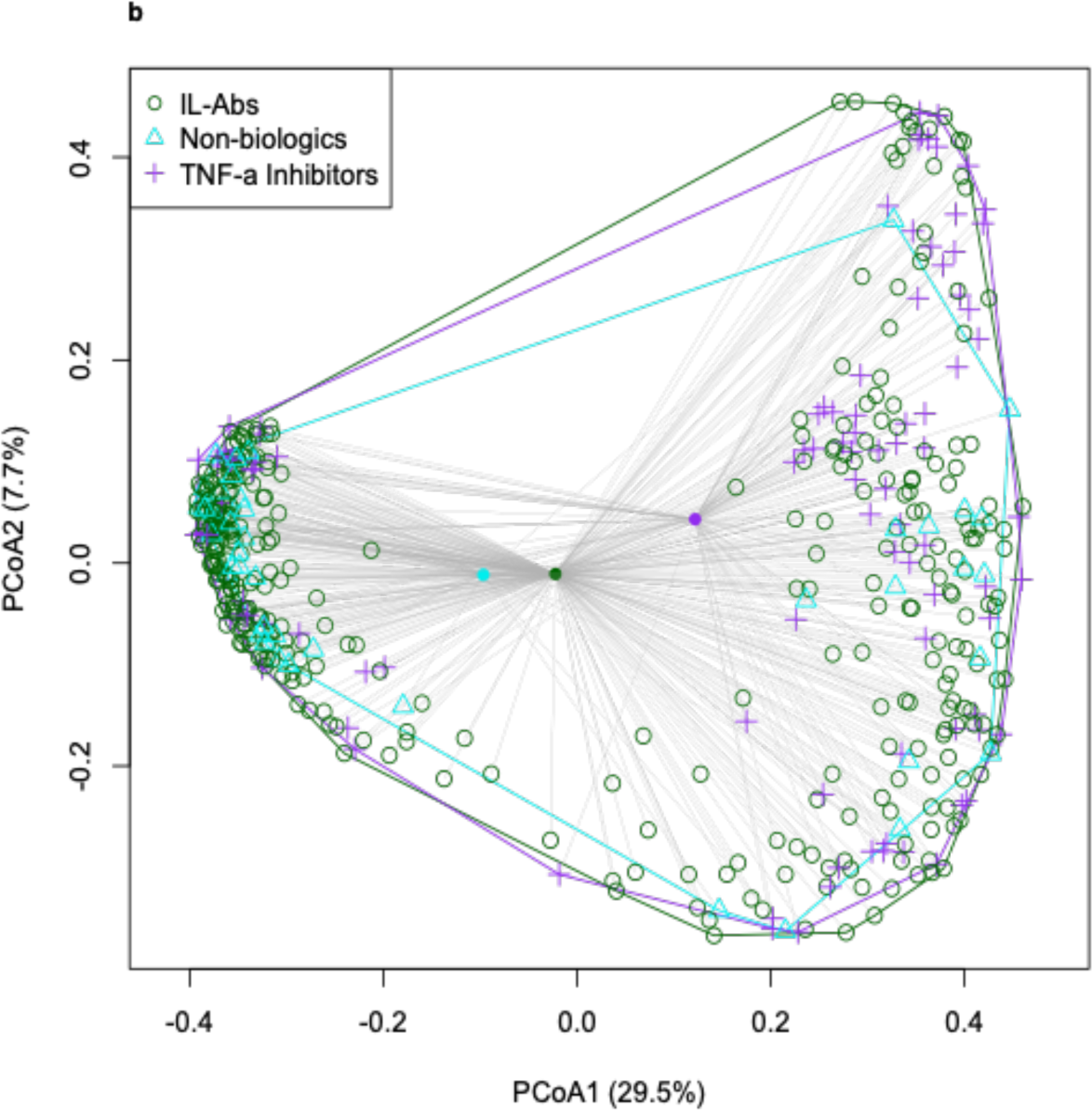
Principal coordinates analysis of genus-level community data from patients of different therapeutic class. Based on 10^4^ permutations, pairwise comparisons of distances between group centroids (solid circles) revealed significant community dissimilarities between TNF-*α* Inhibitor and anti-ILAb patients (P <0.001) or nonbiologics patients (P < 0.001) as well as between non-biologics and anti-ILAb users (P <0.01).

**Table 4.**
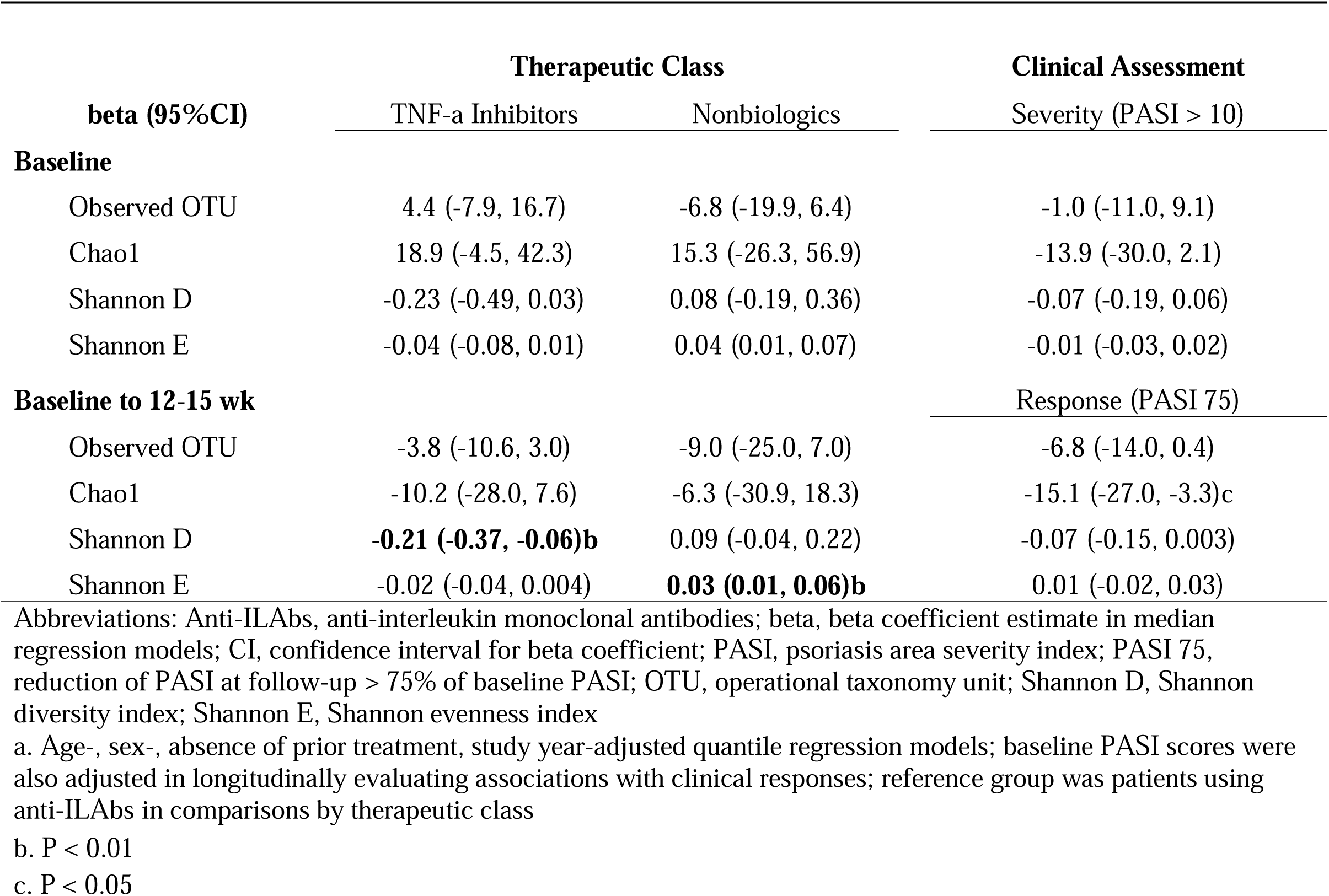
Adjusted associations between genus-level alpha-diversity measures of lesional microbiota and therapeutic class or clinical assessment in adult psoriasis patients based at baseline and longitudinally, 2015-2019^a^.

### Clonal shifts of TCR during treatment

In TCR analysis, we included sequencing results of 8 samples with 4157 (amino acid sequence) unique CDR3 clonotypes (IQR: 2282 - 5642) identified per sample. The average D50 value was 2.8% (IQR: 2.1%-3.2%), suggesting that the most abundant 131 clonotypes accounted for 50% of the total sample reads; there were no statistical differences in D50 values by disease status at baseline (median difference: 0.9%, 95%CI: -4.5%, 6.3%). Shannon Diversity index of TCR repertoire in psoriasis patients varied little after treatment (Table S6). We also showed that, among 10 out of 13 J-and 48 V-segment genes, treatment-associated changes were significant for segment hTRBJ1-6 (+0.9%, 95%CI: 0.4%, 1.4%), hTRBJ2-6 (+1.4%, 95%CI: 0.3%, 2.5%), and hTRBV3-1 (-0.7%, 95%CI: -1.4%, -0.03%) in after-versus before-treatment comparisons among psoriasis patients (Fig. 2).

**Fig 2.**
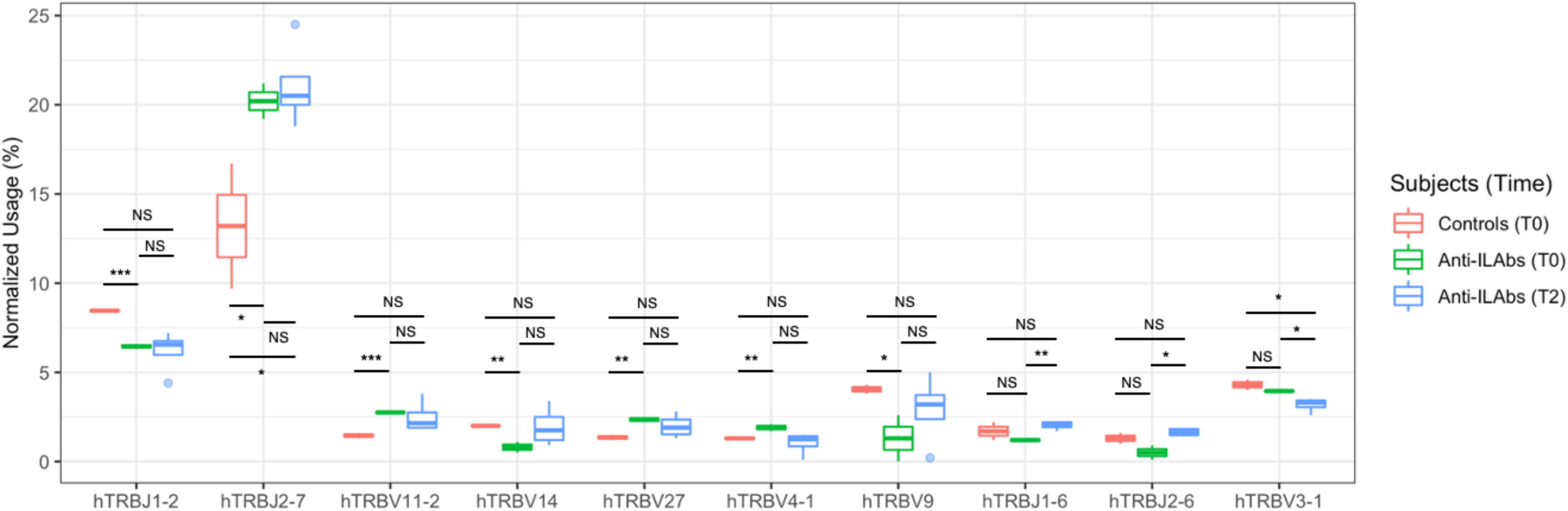
Boxplots of selected hTRB-V and -J segments with significant differential usage in peripheral CD3+ T-cells between skin-healthy controls and psoriasis patients at baseline (T0) and those at 12-15 weeks after anti-ILAb treatment began (T2). Normalized usage was the proportion of each segment being identified in all CDR3 clonotypes of a given sample; the copy number of each clonotype was not taken into account in order to avoid the influence of clonal expansion on estimating sample-specific segment usage. Usage of segment hTRBJ1-2 (difference: 2.2%, 95%CI: 1.8%, 2.6%) and hTRBV9 (difference: 4.3%, 95%CI: 1.0%, 7.6%) were significantly higher in healthy controls than in psoriasis patients before treatment. In contrast, the normalized usage of hTRBJ2-7 (difference: 11.5%, 95%CI: 2.1%, 20.9%), hTRBV11-2 (difference: 1.5%, 95%CI: 1.1%, 1.9%), and hTRBV27 (difference: 1.3%, 95%CI: 0.8%, 1.8%) were statistically higher in psoriasis patients than in controls. NS, non-significant; *: P< .05; **: P< .01; ***: P< .001.

## DISCUSSION

In this prospective cohort study, we showed that, transcriptionally-active skin microbiota in psoriasis patients of 3 major therapeutic classes displayed distinctive community abundances and diversities before and after treatment. Over the past decade, the well-debated observations that TNF-*α* inhibitor could increase patients’ risks for severe infections [28, 29], such as reactivation of latent tuberculosis [30, 31], and the less-debated low risks of similar adverse events in patients receiving IL-17A or IL-12/23p40 inhibitor [32, 33], have led to our hypothesized differential effects of systemic therapies on lesional microbiota. Since both biologics lead to a common, downregulated secretion of cytokines, chemokines, and AMPs, such *β*-defensins and S100 proteins [12], we anticipated starker contrasts between lesional microbiota in biologics and nonbiologics users than between samples from patients receiving different biologics. The observation that only results of *β*-analysis conformed with our conjectures, alluded the multi-dimensional complexity in quantifying (dis)similarities among groups of surveyed human microbiota [21].

Besides specific immune perturbations inflicted on lesional microbiota by different therapies, the initial community structure may partially account for the group differences observed longitudinally. For instance, non-biologics users had the highest relative abundance of Firmicutes (25.8%, IQR: 22.1%, 29.4%) at baseline (anti-ILAb: 20.3%; TNF-*α* inhibitor: 21.8%), yet their lesional microbiota also followed a significant decline temporally (in Firmicutes, - 0.06% per day, 95%CI: -0.09%, -0.02%). Rates of changes in the other 2 treatment groups remained nonsignificant, leading to a consensus post-treatment state across the comparison groups. Contrary to the other two groups, samples from anti-ILAb patients showed evident stability in community abundances, richness, and diversity over the treatment period, rendering the group as an optimal reference for within-patient comparisons.

Our findings that the relative ranking in normalized usage of TCR segments TRBV3-1, V9, and V11-2 examined in the current study, agreed with an early investigation by Tsuda et al. in evaluating effects of ustekinumab on the presence or absence of TRB V-segments in psoriasis patients [34]. These transcriptional shifts in selective CDR3 clonotypes (31% of 13 J-segments, 13% of 48 V-segments) of circulating T cells in patients receiving IL-12/-23 or IL-17A monoclonal antibodies suggested host systemic responses in T cell differentiation after 12-week exposure to blockade against the Th1/Th17 axis. Whether these transitions in TCR clonality resulted from direct depletion of associated lineages or indirect responses to a re-shaping of skin microbiota, was beyond the study’s scope.

Conversely, the notion that characteristics of lesional microbiota were not linked to pre-treatment disease severity, was contrary to a previously-reported negative correlation found between baseline absolute PASI score and Shannon Diversity. In a pre-post study on effects of UVB treatment, Assarsson and colleagues found that there were no similarly positive relationships between disease severity and microbial diversity after treatment [35]. Although *Corynebacterium* sp. has been implicated in IL-1*β* production and expansion of *γδ*_17_ cells in inflammatory skin conditions of model animals [36], our finding that a minor decrease in *Corynebacterium* sp. Was associated with clinical responses during treatment, warrants additional research to confirm its potential clinical utility.

Although the observation that normal skin harbored fewer Proteobacteria than lesional microbiota, was inconsistent with previous findings [13, 37], such cross-study inconsistency has been well-recognized in the literature [27]. Potential sources of disagreements may include heterogeneous patient characteristics under different investigations; choices of analytic approaches; intrinsic discrepancies in DNA-vs. RNA-based profiling of bacterial 16S rRNA genes of skin microbiota. Particularly, Belheouane and others compared the murine ear skin microbiota of standing and active microbial communities using 16S rRNA genes and transcripts sequence data, respectively, and concluded that, RNA-was better than DNA-based profiling in reducing environmental contaminations [6]. Given that comparisons in microbial abundances and a diversity measures between DNA-and RNA-based methods may not always agree with each other even within the same study [8, 14, 38, 39], between-study heterogeneities can be expected; nonetheless, comparative results obtained by either technique shall remain valid within each study [14].

### Limitations

A few limitations are noteworthy in the current analysis. First, as an observational study, we did not specify group sizes for each treatment group *a priori*; restrictions on funding also precluded extended recruitment. The impacts of unequal group sizes and the broad-ranging grouping of patients receiving conventional systemics or anti-ILAb, were uncertainties and possibly attenuated associations. As such, results of comparisons by therapeutic class required careful interpretation and generalization. According to a population-based patient registry in Sweden, female patients generally had a lower PASI score than male patients across the age groups [40]. Since the current study sought to enroll patients with moderate-to-severe psoriasis, the predominance of men in our patient population was expected and the sex-adjusted comparison results within the patient population remained relevant and valid, particularly to those receiving systemic treatment. Although proportions of patients receiving different systemic therapeutics did not differ substantially across the recruitment waves, microbial structure, and diversity of lesional microbiota may be conditioned by patients’ prior treatment, of which we used a proxy variable (the absence of prior treatment) for statistical adjustment; the reported associations were indeed weighted averages of a heterogeneous patient population. Lastly, we recognized the challenges of low biomass even on skin lesions and attempted to sequence cDNA sequences of 16S amplicons instead of the corresponding DNA sequences directly. How this unique approach could have resulted in biased representations of certain Phyla has not been well characterized [41]. Future studies that collect detailed information on serial medications, may provide novel insights on the long-term trajectories of therapeutic effects on the lesional microbiota.

## CONCLUSIONS

The current study showed that, classes of systemic therapies were independently associated with characteristics of lesional microbiota in adult psoriasis patients enrolled in a non-trial setting.

These findings may support for the clinical utility of transcriptionally-active skin microbiota as biomarkers for tailoring systemic therapies to patients with moderate to severe psoriasis in addition to the absolute PASI scores.

## AUTHOR CONTRIBUTIONS

Dr. S Liu had full access to all the data in the study and takes responsibility for the integrity of the data and accuracy of the reported findings.

*Concept and design:* S Liu, YH Huang, T Tsai

*Acquisition, analysis, and interpretation of data:* S Liu, YH Huang, H Weng, T Tsai, H Yu, YC Chiu

*Drafting of the manuscript:* S Liu, H Weng

*Critical revision of the manuscript for important intellectual content:* S Liu, H Weng, T Tsai, YH Huang

*Statistical analysis:* S Liu

*Administrative, technical, or material support:* HY Yang, L Chen, YL Chiu, HY Yu, YC Chiu, C Ng, Y Chang, C Hui, YC Huang

*Supervision:* S Liu

## Data Availability

All data produced in the present study are available upon reasonable request to the authors.

