## Supplemental file for "Temporal dynamics of skin microbiota and immune correlates in psoriasis patients receiving systemic treatment"

**Content:**

Tables S1-S6

Fig. S1-S4

| **Table S1.** Baseline characteristics of psoriasis patients included versus those excluded from the 16S rRNA transcript analysis | | | |
| --- | --- | --- | --- |
| **Characteristics, n (%)** | **Included (N=61)** | **Excluded (N=5)** | P-value^a^ |
| Follow-up time (days), Median (IQR) | 105 (84-112) | 104 (93-104) | 0.429 |
| Age (years), Median (IQR) | 43 (35-50) | 41 (37-41) | 0.390 |
| 20-36 | 31% | 0 | 0.255 |
| 36-50 | 43% | 80% |  |
| ≥ 50 | 26% | 20% |  |
| Male | 87% | 80% | 0.531 |
| BMI (Kg/m^2^), Median (IQR) | 27.8 (26.0-29.8) | 28.1 (23.7-29.9) | 0.592 |
| WC (cm), Median (IQR) | 95 (89-101) | 86 (86-92) | 0.352 |
| Education |  |  |  |
| Less than high school | 30% | 0 | 0.425 |
| High or vocational school | 36% | 60% |  |
| College or higher | 34% | 40% |  |
| Disease duration (years), Median (IQR) | 23 (19-33) | 30 (26-32) | 0.352 |
| Treatment-naive | 30% | 20% | 1.000 |
| Regular alcohol use | 21% | 0 | 0.574 |
| Regular smoking | N=57 |  |  |
| Never | 57% | 60% | 0.600 |
| Quitted | 8% | 20% |  |
| Yes | 34% | 20% |  |
| Regular use of antiseptics for |  |  |  |
| Face | 5% | 20% | 0.276 |
| Hand^b^ | 8% | 25% | 0.328 |
| Staphylococcal colonization |  |  |  |
| Lesional skin | 31% | 40% | 0.650 |
| Nares | 21% | 20% | 1.000 |
| Therapeutic class |  |  |  |
| Non-biologics | 10% | 60% | 0.015 |
| Biologics |  |  |  |
| TNF-a Inhibitors | 21% | 20% |  |
| Anti-ILAbs | 69% | 20% |  |
| Clinical outcomes |  |  |  |
| PASI, Median (IQR) | 15 (11-20) | 3 (3-16) | 0.642 |
| DLQI, Median (IQR) | 9 (4-15) | 12 (4-12) | 0.642 |
| CESD score, Median (IQR) | 16 (12-22) | 17 (13-19) | 0.592 |
| Abbreviations: BMI, body mass index; CESD, Center for Epidemiologic Studies Depression Scale; DLQI, Dermatology Life of Quality Index; IQR, interquartile range; PASI, psoriasis area severity index; WC, waist circumference | | | |
| a. P-value for median test; Chi-square test, or Fisher's exact test | | | |

| **Table S2.** Temporal changes in patient-important clinical outcomes in psoriasis patients overall and by therapeutic class, 2015-2019 | | | | | | |
| --- | --- | --- | --- | --- | --- | --- |
| Median (IQR) | **All (N=61)** | P for trend^a^ | **Therapeutic Class** | | | |
|  |  |  | Non-biologics (N=6) | TNF-𝛼 inhibitors (N=13) | Anti-ILAbs (N=42) | P for differential trend^b^ |
| **PASI** |  |  |  |  |  |  |
| Baseline | 15 (11-20) | <0.001 | 15 (14-20) | 13 (11-17) | 15 (11-24) | 0.197 |
| 4-6 Week | 7 (5-12) |  | 7 (4-48) | 8 (8-10) | 7 (5-12) |  |
| 12-15 Week | 4 (1-8) |  | 9 (9-12) | 5 (2-9) | 3 (1-6) |  |
| 18-25 Week | 5 (3-12) |  | 5 | 5 (2-11) | 8 (3-25) |  |
| 28-29 Week | 4 (0-9) |  | - | 0 | 8 (0-11) |  |
| 37-42 Week | 4 (2-9) |  | 2 | 4 (2-9) | 6 (4-9) |  |
| 48-57 Week | 5 (0-9) |  | 2 | 3 (0-8) | 8 (3-10) |  |
| **DLQI** |  |  |  |  |  |  |
| Baseline | 9 (4-15) | <0.001 | 12 (8-22) | 9 (4-14) | 10 (3-15) | 0.565 |
| 4-6 Week | 10 (3-14) |  | 4 (2-23) | 10 (3-15) | 8 (3-14) |  |
| 12-15 Week | 2 (0-8) |  | 10 (4-13) | 2 (0-8) | 2 (0-7) |  |
| 18-25 Week | 3 (1-10) |  | 3 | 1 (0-7) | 9 (5-14) |  |
| 28-29 Week | 6 (2-10) |  | - | 0 | 8 (3-11) |  |
| 37-42 Week | 3 (0-9) |  | 4 | 0 (0-12) | 3 (0-9) |  |
| 48-57 Week | 2 (0-6) |  | 2 | 0 (0-5) | 4 (1-8) |  |
| **CESD** |  |  |  |  |  |  |
| Baseline | 16 (12-22) | 0.234 | 17 (12-24) | 13 (11-20) | 16 (13-22) | 0.960 |
| 4-6 Week | 14 (11-20) |  | 14 (4-28) | 11 (10-20) | 14 (11-19) |  |
| 12-15 Week | 15 (12-19) |  | 21 (21-23) | 13 (12-18) | 15 (12-18) |  |
| 18-25 Week | 15 (14-21) |  | 9 | 15 (13-18) | 18 (15-24) |  |
| 28-29 Week | 17 (12-22) |  | - | 27 | 17 (13-22) |  |
| 37-42 Week | 13 (10-18) |  | 4 | 12 (9-20) | 18 (17-20) |  |
| 48-57 Week | 16 (13-18) |  | 4 | 13 (12-17) | 18 (14-18) |  |
| Abbreviations: CESD, Center for Epidemiologic Studies Depression Scale; DLQI, Dermatology Life of Quality Index; PASI, psoriasis area severity index | | | | | | |
| a. P-value for temporal trend from baseline to week 12-15, adjusting for age, sex, absence of prior treatment, and cohort year in median regression models with robust variance estimation to account for repeated measurements | | | | | | |
| b. P-value for differential effects by therapeutic class on temporal trends (within 12-15 weeks of treatment) using multivariable median regression models and robust variance estimation method | | | | | | |

| **Table S3.** Differences in genus-level relative abundance between lesional and healthy skin microbiota at baseline and during 12-15 weeks of systemic treatment, 2016-2019^a^ | | | | | |
| --- | --- | --- | --- | --- | --- |
| Relative | **Baseline** | |  | **Baseline to 12-15 wk** | |
| Abundances (%) | Unadjusted | Adjusted^a^ |  | Unadjusted | Adjusted^a^ |
| **Genus** | beta (95%CI) | beta (95%CI) |  | beta (95%CI) | beta (95%CI) |
| Corynebacterium | 0.27 (-0.15, 0.70) | -0.01 (-0.34, 0.32) |  | -0.05 (-0.33, 0.23) | -0.08 (-0.27, 0.11) |
| Propionebacterium | -0.15 (-0.44, 0.13) | -0.09 (-0.73, 0.55) |  | 0.08 (-0.11, 0.27) | -0.12 (-0.39, 0.15) |
| Staphyloccous | -0.43 (-2.15, 2.19) | -0.24 (-2.19, 1.70) |  | -0.36 (-1.49, 0.77) | -0.08 (-1.25, 1.08) |
| Streptococcus | 0.13 (-0.60, 0.85) | -0.19 (-1.20, 0.83) |  | -0.09 (-0.59, 0.42) | 0.15 (-0.52, 0.82) |
| Veillonella | -0.02 (-0.11, 0.08) | 0.17 (-0.10, 0.44) |  | 0.03 (-0.04, 0.10) | -0.09 (-0.25, 0.08) |
| Enhydrobacter | 0.07 (-0.10, 0.24) | -0.02 (-0.33, 0.28) |  | -0.03 (-0.14, 0.09) | -0.07 (-0.22, 0.09) |
| Abbreviations: beta, beta coefficient estimate in median regression models; CI, confidence interval for beta coefficient | | | | | |
| a. Age-, sex-, absence of prior treatment, skin microenvironment, study year-adjusted in baseline models; follow-up duration was also accounted for in the longitudinal models; all comparisons excluded data from 2015-2016, during which no control subjects were enrolled | | | | | |
| b. P < 0.01 |  |  |  |  |  |
| c. P < 0.05 |  |  |  |  |  |
| d. P ≤ 0.001 |  |  |  |  |  |

| **Table S4.** Results of multivariable quantile regression analysis on alpha-diversity metrics comparing lesional to normal skin microbiota at baseline and over the study period, 2016-2019 | | | |
| --- | --- | --- | --- |
|  | **beta** | **(95%LL, UL)** | P-value |
| **Baseline** |  |  |  |
| Observed OTU | 0 | (-9.0, 9.0) | 1.000 |
| Chao1 | -5.6 | (-0.9, 0.1) | 0.477 |
| Shannon D | -0.05 | (-0.20, 0.10) | 0.528 |
| Shannon E | **-0.02** | **(-0.03, -0.005)** | **0.010** |
| **Baseline to 12-15 wk** |  |  |  |
| Observed OTU | 1.0 | (-3.8, 5.8) | 0.680 |
| Chao1 | 0.50 | (-9.7, 10.7) | 0.921 |
| Shannon D | -0.01 | (-0.09, 0.08) | 0.899 |
| Shannon E | -0.01 | (-0.02, 0.01) | 0.349 |
| Abbreviations: beta, beta coefficient estimate in median regression models; CI, confidence interval for beta coefficient; OTU, operational taxonomy unit; Shannon D, Shannon diversity index; Shannon E, Shannon evenness index | | | |
| a. Age-, sex-, absence of prior treatment, study year-adjusted quantile regression models; follow-up intervals were also added to the longitudinal model; these comparisons exclude data from year 2015-2016 | | | |

| **Table S5.** Adjusted associations between genus-level relative abundances of lesional microbiota and therapeutic class or clinical outcome at baseline and over time in psoriasis patients, 2015-2019^a^ | | | | |
| --- | --- | --- | --- | --- |
| Relative Abundance (%) | **Therapeutic Class** | |  | **Clinical Assessment** |
|  | TNF-a | Non-biologics |  | Severity (PASI > 10) |
| **Baseline** | beta (95%CI) | beta (95%CI) |  | beta (95%CI) |
| Corynebacterium | -0.46 (-0.98, 0.07) | -0.30 (-0.90, 0.31) |  | -0.18 (-0.70, 0.35) |
| Propionebacterium | -0.02 (-0.10, 0.06) | -0.02 (-0.34, 0.29) |  | 0.05 (-0.01, 0.11) |
| Staphyloccous | 1.00 (-0.55, 2.54) | **5.90 (2.36, 9.43)b** |  | -0.23 (-1.75, 1.28) |
| Streptococcus | 0.25 (-0.83, 1.33) | -0.24 (-1.42, 0.94) |  | -0.30 (-1.17, 0.57) |
| Veillonella | -0.00 (-0.04, 0.04) | 0.09 (-0.05, 0.23) |  | -0.01 (-0.17, 0.15) |
| Enhydrobacter | -0.09 (-0.30, 0.12) | 0.08 (-0.14, 0.29) |  | -0.31 (-0.65, 0.02) |
| **Baseline to 12-15 wk** |  |  |  | Response (PASI 75) |
| Corynebacterium | -0.13 (-0.38, 0.12) | -0.01 (-0.39, 0.38) |  | **-0.38 (-0.65, 0.12)c** |
| Propionebacterium | -0.02 (-0.07, 0.03) | 0.18 (-0.23, 0.59) |  | 0.01 (-0.08, 0.11) |
| Staphyloccous | 1.07 (-0.44, 2.58) | 1.49 (-0.59, 3.58) |  | -0.63 (-2.07, 0.81) |
| Streptococcus | -0.35 (-0.73, 0.02) | 0.43 (-0.44, 1.30) |  | -0.15 (-0.56, 0.27) |
| Veillonella | -0.002 (-0.04, 0.03) | 0.04 (-0.004, 0.09) |  | -0.01 (-0.08, 0.05) |
| Enhydrobacter | -0.00 (-0.21, 0.20) | 0.14 (-0.02, 0.30) |  | -0.01 (-0.20, 0.18) |
| Abbreviations: Anti-ILAbs, anti-interleukin monoclonal antibodies; beta, beta coefficient estimate in median regression models; CI, confidence interval for beta coefficient; PASI, psoriasis area severity index | | | | |
| a. Age-, sex-, absence of prior treatment, study year, abd follow-up duration-adjusted quantile regression models; baseline PASI scores were also adjusted in evaluating associations with clinical responses; reference group was patients using anti-ILAbs in comparisons by therapeutic class | | | | |
| b. P ≤ 0.001 |  |  |  |  |
| c. P < 0.01 |  |  |  |  |

| **Table S6.** Results of TCR profiling in selected controls (N=7) and patients receiving biologics treatment (N=9) | | | | | | |
| --- | --- | --- | --- | --- | --- | --- |
|  | **Total** | **Control** | | **Psoriasis Patients** | | |
| Sampling time |  | Baseline | | Baseline | | 12-15 Week |
| No. patients | 16 | 7 | | 6 | | 9 |
| No. pooled blood samples | 8 | 2 | | 2 | | 4 |
| TCR Diversity Metrics |  |  | |  | |  |
| D50 no. | 131 (30-167) | 112 (40-185) | | 117 (16-217) | | 131 (65-151) |
| D50 value | 2.8 (2.1-3.2) | 2.1 (1.1-3.1) | | 3.1 (2.1-4) | | 2.8 (2.4-3.1) |
| Diversity index | 8.5 (7.1-8.5) | 8.7 (6-8.7) | | 8.6 (4.4-8.6) | | 8 (8-8.5) |
| Shannon Diversity | 9.3 (7.2-9.7) | 8.9 (8-9.8) | | 8.0 (6-9.9) | | 9.3 (7.8-9.5) |
| Segment usage, normalized % | | |  | |  | |
| J segment, median (IQR) | **6.2 (2-12.1)** | **6.6 (2.1-13.2)** | | **5.8 (1.8-11.9)** | | **5.7 (2-11.7)** |
| hTRBJ1-2 | 6.6 (6.4-7.8) | 8.5 (8.4-8.5) | | 6.5 (6.3-6.6)^a^ | | 6.6 (5.5-6.9) |
| hTRBJ1-6 | 1.9 (1.3-2.2) | 1.7 (1.2-2.2) | | 1.2 (1.1-1.3) | | 2.1 (1.9-2.2)^b^ |
| hTRBJ2-6 | 1.5 (1.0-1.7) | 1.3 (1-1.6) | | 0.5 (0.1-0.9) | | 1.7 (1.5-1.9)^b^ |
| hTRBJ2-7 | 19.8 (17.8-20.9) | 13.2 (9.7-16.7) | | 20.2 (19.2-21.2)^c^ | | 20.5 (19.6-22.6) |
| V segment, median (IQR) | **1.1 (0.1-2.8)** | **1.1 (0.2-2.8)** | | **1.0 (0-2.8)** | | **1.1 (0.1-2.8)** |
| hTRBV3-1 | 3.7 (3.3-4) | 4.3 (4-4.6) | | 4.0 (3.9-4) | | 3.3 (2.9-3.5)^bc^ |
| hTRBV4-1 | 1.4 (1.2-1.6) | 1.3 (1.2-1.4) | | 1.9 (1.7-2.1)^c^ | | 1.3 (0.6-1.5) |
| hTRBV9 | 3.2 (1.4-4.1) | 4.1 (3.8-4.3) | | 1.3 (0-2.6)^c^ | | 3.2 (1.7-4.2) |
| hTRBV11-2 | 2.2 (1.8-2.8) | 1.5 (1.3-1.6) | | 2.8 (2.7-2.8)^a^ | | 2.2 (1.9-3.1) |
| hTRBV14 | 1.7 (1-2.4) | 2 (2-2) | | 0.8 (0.5-1.1)^d^ | | 1.8 (1.1-2.8) |
| hTRBV27 | 1.9 (1.4-2.5) | 1.4 (1.2-1.5) | | 2.4 (2.2-2.5)^d^ | | 1.9 (1.5-2.5) |
| Abbreviations: D50 no., cumulative number of unique clonotypes that cumulatively comprise 50% of all sequence reads in a given sample; D50 value, propotion of D50 no. out of total unique number of clonotypes in a given sample; Di, Diversity Index; IQR, interquartile range; K, 1000; No., number; TCR, T cell receptor; CDR, Complementarity-determining region | | | | | | |
| a. P < 0.001 for median comparison with controls at baseline using bootstrapped quantile regression | | | | | | |
| b. P < 0.05 for median comparison with patients at baseline using bootstrapped quantile regression | | | | | | |
| c. P < 0.05 for median comparison with controls at baseline using bootstrapped quantile regression | | | | | | |
| d. P < 0.01 for median comparison with controls at baseline using bootstrapped quantile regression | | | | | | |

**Fig. S1.** Participant and sample flow chart. Between 2015 and 2019, we recruited and followed 119 participants for at least 12 weeks. After excluding 24 ineligible participants, we followed 66 psoriasis patients and 29 skin-healthy participants, among whom 95 contributed 1182 skin and nasal swabs for 16S rRNA gene sequencing. We included 743 and 226 swabs from 61 psoriasis and 29 skin-healthy controls in the current analysis after excluding low-quality sequencing samples (< 400 reads/ sample).

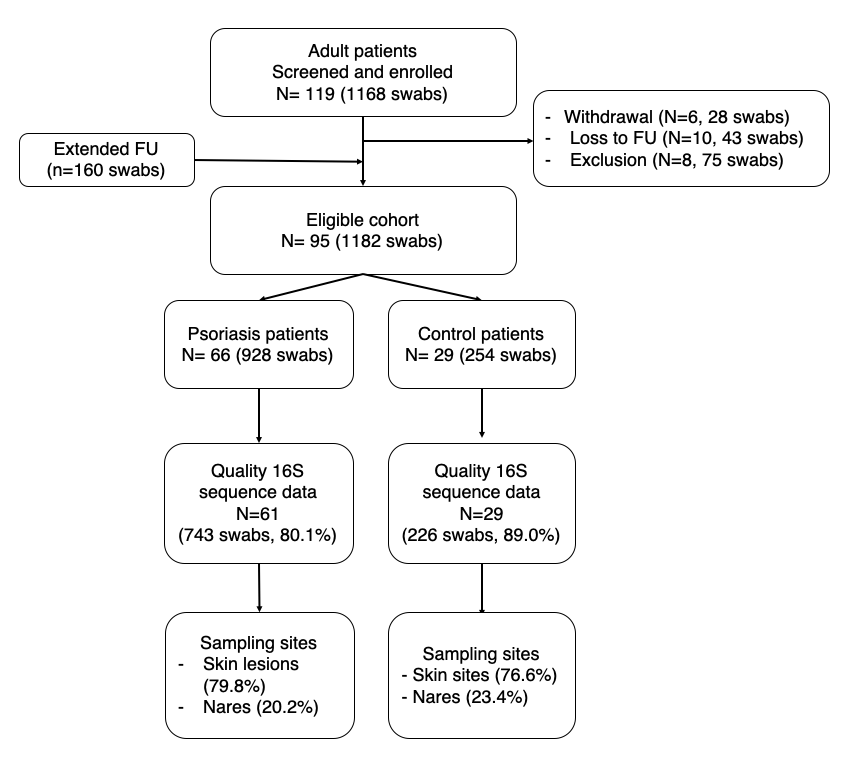

**Fig. S2a.** Prior to treatment began (Day 0), there were no significant differential phylum abundances across the four groups. Over time, a relative over-abundance of Proteobacteria was found in psoriatic lesions as compared to normal skin (3.35%, 95%CI: 1.44%, 5.25%) with adjustment for potential confounding.

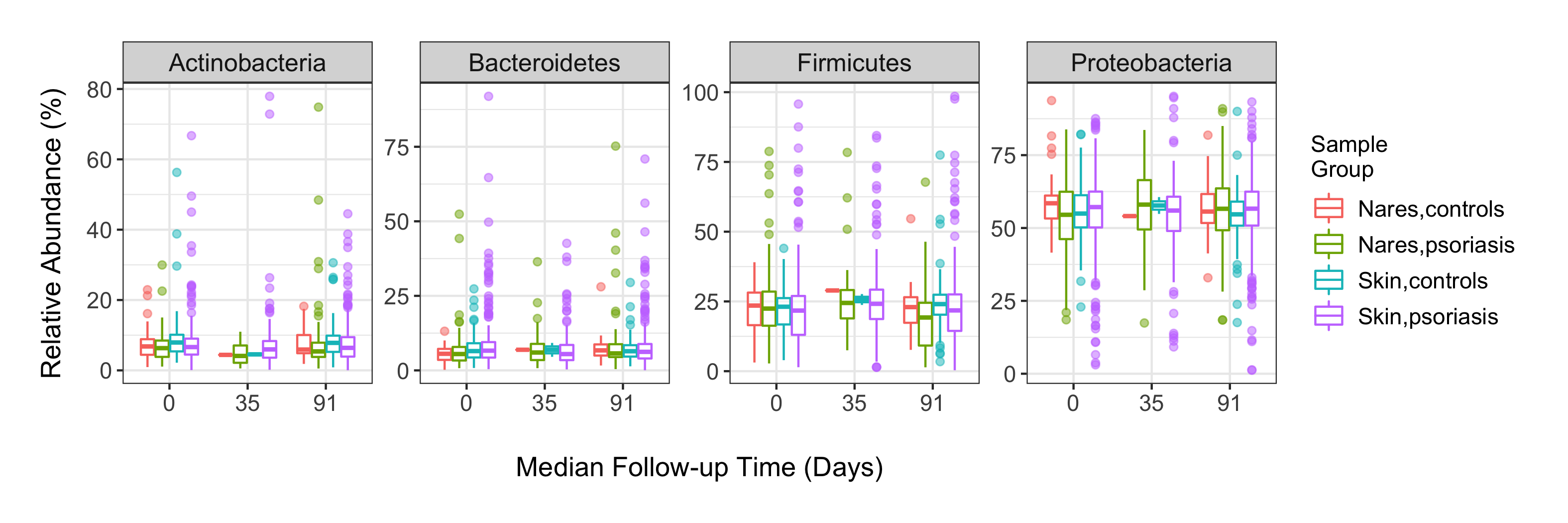

**Fig. S2b.** Relative abundances of selected genera in skin microbiota grouped by disease status (control vs. patient) and sampling site (nonlesional vs. lesional). There were no significant indicator taxa across the comparison groups. We did not include *Micrococcus* sp. due to its absence in some samples.

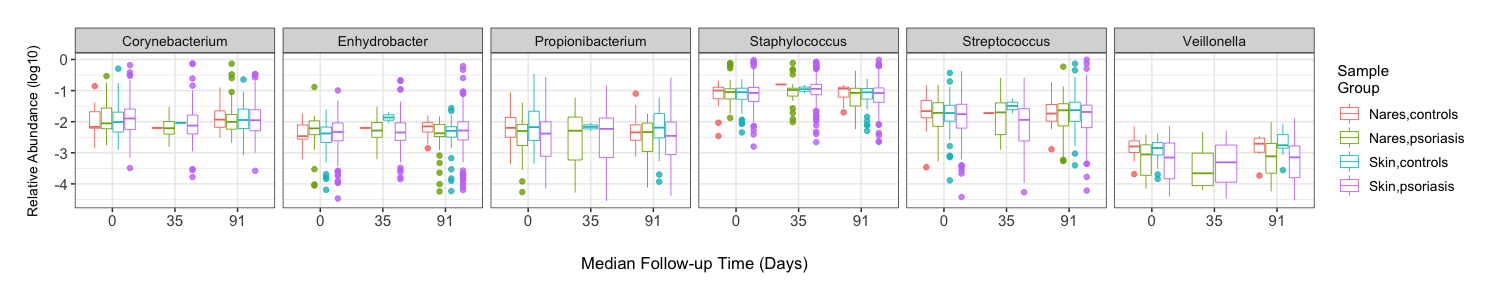

**Fig. S2c.** Principal coordinates analysis of community data from skin samples of healthy controls and psoriasis patients based on Bray-Curtis dissimilarity. Results of hypothesis testing using adonis function (R package: vegan) were not statistically significant (P=0.638) based on 10 000 permutations.

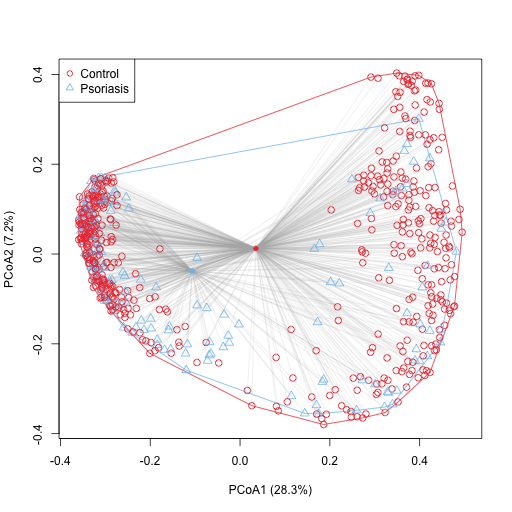

**Fig. S3.** Relative abundances of major phyla in skin microbiota of skin-healthy participants and psoriasis patients by therapeutic class during the study period. Baseline lesional microbiota of anti-ILAb and TNF-𝛼 Inhibitor users showed a 5.74% higher and 1.68% lower abundance of Proteobacteria (95%CI: 2.33%, 9.16%) and Actinobacteria (95%CI: -2.90%, -0.46%) than normal skin microbiota, separately. The average difference between TNF-𝛼 Inhibitor users and skin-healthy individuals remained throughout the treatment period (-1.68%, 95%CI: -2.75%, -0.61%) whereas those differences between patients receiving TNF-𝛼 Inhibitor and anti-ILAb attenuated.

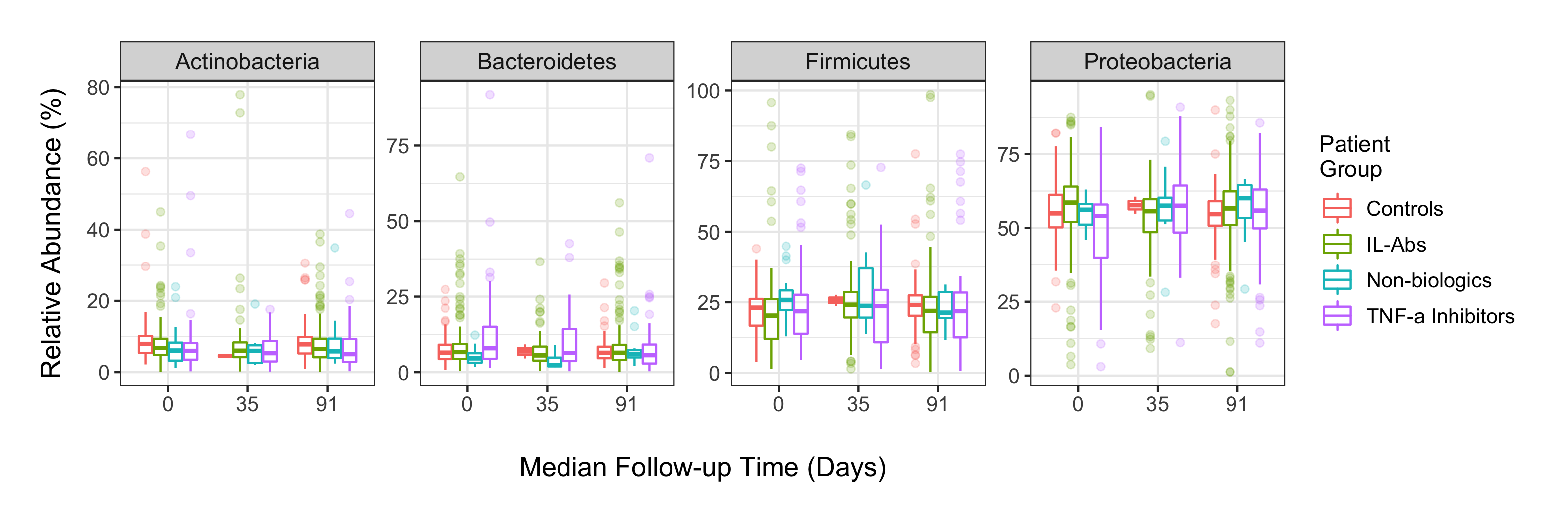

**Fig. S4.** Principal coordinates analysis of community data from skin-healthy controls and psoriasis patients by (a) therapeutic class and (b) clinical PASI 75 responses. Solid dots are group centroids. (a) Results of four-group comparisons suggested significant distances among the four group centroids (permuted P < 0.001); pairwise comparisons further showed uniform differences between patients and healthy controls, except for the nonbiologics group (permuted P=0.444). The discrepant results between Fig. S4a and Fig. S2c indicated that the two-group comparison was confounded (biased). (b) Pairwise comparisons between skin microbiota from control participants and PASI 75 responders or non-responders were statistically significant (both permuted P<0.001). In contrast, within the patient group, there was no discernable ecological difference between good and poor responders (permuted P=0.090).

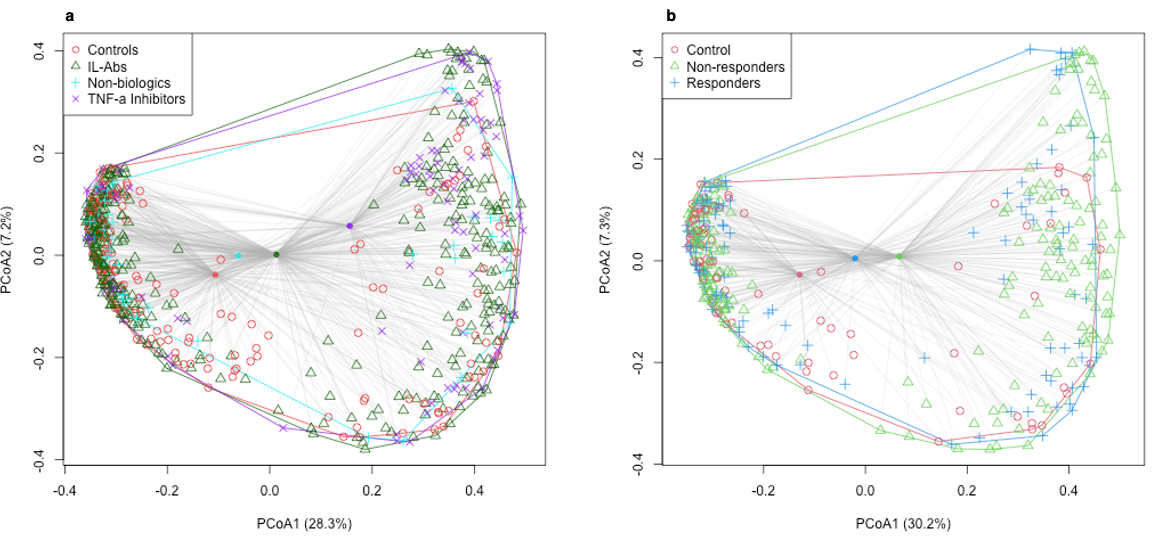
